# Transcriptional alteration in TRKβ-SHC isoform as a neuroprotective factor for post stroke memory outcome

**DOI:** 10.64898/2026.02.21.26346768

**Authors:** Dipanwita Sadhukhan, Raktim Choudhury, Saranya Roy, Arunima Roy, Shreyosi Maitra, Kartik Chandra Ghosh, Joydeep Mukherjee, Tapas Kumar Banerjee, Subhra Prakash Hui, Saikat Chakrabarti, Arindam Biswas

## Abstract

**Background:** Post-stroke cognitive impairment (PSCI) affects nearly 30% of stroke survivors and significantly impairs functional recovery. Brain-derived neurotrophic factor (BDNF)–tropomyosin receptor kinase-β (Trkβ) signalling is crucial for synaptic plasticity and cognitive function. While altered expression of truncated TRKβ-T1 isoforms has been linked to stroke, the contribution of the TRKβ-SHC isoform to PSCI in humans remains poorly understood.

**Objectives:** This study aimed to (i) assess isoform-specific expression changes of NTRK2 associated with PSCI, (ii) evaluate the role of an isoform-specific genetic variant in disease susceptibility, and (iii) identify DNA methylation changes regulating NTRK2 expression (if any).

**Methods:** Gene expression levels of three major NTRK2 isoforms and MEK2 were analyzed in peripheral blood mononuclear cells from 19 PSCI patients, 21 post-stroke cognitively normal (PSCN) individuals, and 11 healthy controls. Expression data were correlated with raw memory scores and MEK2 expression. DNA methylation profiles of NTRK2 and its transcriptional regulators were assessed using whole-genome bisulfite sequencing.

**Results:** TRKβ-FL expression was significantly reduced in stroke patients compared with controls. In contrast, TRKβ-SHC expression was elevated in PSCN individuals relative to PSCI cases and showed a positive correlation with MEK2 expression and memory performance. No significant association was observed between rs65339833 and cognitive subdomains. Gene body hypermethylation, but not promoter methylation, was detected in NTRK2 and its regulatory genes.

**Conclusions:** Elevated TRKβ-SHC expression may contribute to preserved cognitive function following stroke. DNA methylation status of NTRK2 may regulate alternative splicing and thus represent a novel therapeutic avenue for preventing or mitigating PSCI.

## Introduction

Stroke is a major cause of death, worsening of daily lifestyle and poor cognition among adults worldwide (Feigin et al. 2022). Among the post stroke survivors, either independent or concomitant onset of cognitive decline (PSCI) or (PSD) depression are major clinical suffering (El Husseini et al. 2023). Several cognitive domains, like attention, memory, language, and orientation, are affected over time. The Post Stroke Cognitive Impairment (PSCI) and Post Stroke Depression (PSD) influence each other and share overlapping disease mechanisms, risk factors, molecular pathways and treatment strategies (Terroni et al. 2012). An increasing number of evidences have established the compromised neuroprotective effects of Brain-derived neurotrophic factor (BDNF) - Tropomyosin-related kinase B receptor (Trkβ) signalling pathway as an underlying mechanism through experimental ischemic stroke (IS) models, and genetic associations of single nucleotide polymorphisms of these genes have been reported in ethnicity dependent manner (Alnoaman et al. 2025, Shi et al. 2020). An alteration in transcript and protein level (in serum of human patients) of BDNF is a common finding for both PSCI and PSD (Sadhukhan et al. 2023, Chang et al. 2024). On the other hand, the TRKβ, the cognate receptor of BDNF, encoded by NTRK2, is only examined as a susceptible gene through population-based study in stroke associated outcomes (Shi et al. 2020).

Like BDNF, the transcription of TRKβ is also complex, and its isoforms show cell specific expression pattern. TRKβ can generate more than 30 isoforms, of which three major splicing isoforms, like TRKβ-FL, TRKβ-SHC, and TRKβ-T1, are well studied. The TRKβ-FL contains a tyrosine kinase domain, an SHC-binding domain, and a PLC-γ-binding domain, while TRKβ-SHC has an SHC-binding domain, and Trkβ-T1 does not have any functional domain. Upon activation by BDNF, TRKβ-FL undergoes dimerisation and autophosphorylation on the intracytoplasmic kinase side, leading to the phospholipase-Cγ (PLCγ) mediated pathways through downstream protein kinase-C signalling and promotes synaptic plasticity. TRKβ-T1 is expressed in both neurons and glia, whereas TRKβ-SHC is neuron-specific (Tessarollo et al. 2022). Its truncated variant *i.e.* TRKβ-T1, whose overexpression is reported in several brain disorders, acts as a dominant negative isoform by competing with BDNF and forming heterodimers with TRKβ-FL (Li et al. 2022, Alsalloum et al. 2023), while the other variant, TRKβ-SHC, appeared as a neuroprotective molecule in an in*-vitro* system by regulating inappropriate BDNF/TRKβ-TK+ signalling after exogenous BDNF treatment (Wong et al. 2012). The altered expression of truncated TRKβ isoforms and imbalance in their expression profile have been identified as the underlying mechanism in a number of neural diseases (Tessarollo et al. 2022).

To this complex transcriptional regulation of TRKβ, it is believed that the small miRNAs could be a potent regulator, suggesting the implication of functional 3’UTR variants in disease pathogenesis (Wong et al. 2014). Likewise, epigenetic regulation of TRKβ was also evaluated in altered TRKβ splicing, which was seen in different neurological disorders (Ernst et al. 2009, Duarte-Ruiz et al. 2025). A previous report describing the isoform-specific differentially methylated sites (DMSs) in regions flanking the unique TRKβ-T1 protein coding sequence (CDS) in CNS-specific cell types also suggested a role of DNA methylation patterns for isoform-specific TRKβ expression (Wei et al. 2024).

Given this background, it is clear that the TRKβ isoform expression profile and examination of regulatory elements for differential splicing (if any) in stroke-related cognitive decline have gained limited emphasis compared to AD (Alzheimer’s Disease) and schizophrenia. Here, we hypothesised that alterations in the abundance of TRKβ isoforms’ mRNA exist in PSCD and may be regulated by cis-regulatory regions and epigenetic modifications. Therefore, we sought to determine the a) expression profile of TRKβ isoforms in PSCI patients with memory impairment, followed by its correlation with relevant secondary messengers’ expression, b) comparative evaluation of differentially methylated sites between PSCI and PSCN patients showing differential isoform expression (if any), and c) genetic susceptibility to PSCI and PSD by 3’UTR SNPs.

## Materials and Methods

### Patients and Controls

The present study recruited a total of 314 post-stroke survivors (age >18 years and neuroimaging evidence of Ischemic stroke) from the National Neurosciences Centre, Calcutta and Nil Ratan Sircar Medical College & Hospital, Kolkata. Informed consent was taken as per the guidelines of the Indian Council of Medical Research (ICMR). All had Bengali as their mother tongue, able to comprehend & speak, and able to read & write. The cases with dysphasia or any other form of language deficit, psychiatric illness, substance abuse, history of psychotropic drug intake, cancers, drugs known to cause cognitive impairment and major head injury were excluded from the study. The demographic details of patients are summarised in Table 1. The history of known risk factors for stroke, including hypertension, diabetes mellitus, hyperlipidemia, and ischemic heart disease, was obtained from the patients’ medical files, while data on alcohol consumption or smoking were collected by asking verbal questions. In addition, age- and ethnicity-matched unrelated healthy controls (n = 11) with no personal or family history of stroke or any neurological symptoms were also recruited in the present study from Kolkata.

**Table 1:** Demographic characteristics of Post-stroke Cognitive Impaired (PSCI), Post-stroke Cognitive Normal (PSCN), and Control subjects.

| Demographic details/subjects | PSCI (n = 152) | PSCN (n = 162) | Controls (n = 11) | P-value |
| --- | --- | --- | --- | --- |
| Age range (years) | 28 - 79 | 28 - 79 | 52-73 | NA |
| Age at onset ± SD | 52.97 ± 12.07 | 49.34±10.55 | NA | **0.03797** |
| Male (n = 254) | 116 (46%) | 138 (54%) | 6 (55%) | **0.0474** |
| Female (n = 60) | 36 (60%) | 24 (40%) | 5 (45%) |  |
| Family History of Stroke (n = 164) | 74 (45%) | 90 (55%) | NA | 0.2235 |
| Hypertension (n = 234) | 110 (47%) | 124 (53%) | 0 (0%) | 0.3966 |
| Diabetes (n = 120) | 64 (53%) | 56 (47%) | 0 (0%) | 0.1701 |
| BMI | 23.09 ± 3.37 | 22.72±2.82 | 23.48 ± 2.42 | 0.7663 |
| NIHSS at admission | 8.13 ± 4.16 | 7.21 ± 4.99 | NA | 0.3029 |
| Arterial involvement, MCA (n = 234) | 104 (45.5%) | 130 (55.5%) | NA | **0.0170** |
| Arterial involvement, PCA (n = 80) | 48 (60%) | 32 (40%) | NA |  |
| Depressed (GDS ≥ 10) | 108 (71%) | 90 (55.56%) | 0 (0%) | **0.0047** |
| Non-Depressed (GDS < 9) | 44 (29%) | 72 (44.44%) | 11 (100%) |  |

PSCI: Post-stroke Cognitive Impaired; PSCN: Post-stroke Cognitive Normal; GDS: Geriatric Depression Scale; NIHSS: National Institute of Health Stroke Scale; MCA: Middle cerebral atrophy; PCA: Posterior cerebral atrophy; BMI: Basal metabolic index. P-value was calculated between PSCI and PSCN cases. NA: Not applicable

### Cognitive Parameters

Cognitive assessment was carried out within 6 to 12 months of stroke onset. The evaluation was done as mentioned in Sadhukhan et al. 2023. In brief, comprehensive cognitive tests for attention, language, memory, visuospatial skills, executive functions, etc., were analysed in detail using Kolkata Cognitive Screening Battery for primary assessment (Biswas et al. 2021). Post-stroke depression (PSD) assessment was done using the Geriatric Depression Scale (GDS) within 6-12 months of stroke onset.

### Selection of SNV

Here, we have genotyped a single-nucleotide variant (SNV) rs6599833T/C located in the 3 - UTR of TRKβ-SHC encoding NTRK2 transcript showing a putative miRNA binding site upon bioinformatic analysis.

### Genomic DNA isolation and genotyping of SNV

Genomic DNA isolation from fresh whole blood was done by the conventional salting out method using sodium perchlorate, followed by isopropanol precipitation (Johns et al. 1989), and then dissolved in TE (10-mM Tris–HCl, 0.1-mM EDTA, pH 8.0). Polymerase chain reaction (PCR) was carried out using the specific primer pairs. Genotypes for SNVs were determined by restriction digestion of their PCR products with suitable enzymes (New England Biolabs Inc., Ipswich) following manufacturers’ protocol. The digested products were separated by 7% polyacrylamide gel electrophoresis and visualised by ethidium bromide staining. About 10% of the samples for each variant were randomly selected for sequencing to rule out genotyping errors.

### Gene Expression Analysis

For the gene expression study, peripheral blood mononuclear cells (PBMCs) were first isolated from fresh whole blood collected from stroke patients during follow up study period and from healthy controls (between 11:30 am and 12 pm to avoid phase differences) using Histopaque 1077 (Sigma-Aldrich, St. Louis, US) double-gradient density centrifugation. Total RNA from (Peripheral Blood Mononuclear Cells) PBMC was extracted using Trizol reagent (Thermo Fisher Scientific, US) and dissolved in DEPC-treated water, stored at −80^0^C. Afterwards, an equal amount of RNA (2 µg) from each sample was subjected to cDNA synthesis using the reverse transcriptase Kit (Promega Corporation, USA) following standard procedures. With these cDNAs, the quantitative real-time PCR (qRT-PCR) was performed to TRKβ-FL, TRβ-SHC, TRKβ-T1, PLCγ1, and MEK1 expression using gene-specific primers designed with the web-based software Primer3 and SYBR Green PCR Master mix (Applied Biosystems, US). While, for normalization purpose, expression of 18S rRNA was used (18S_F: 5’-GTAACCCGTTGAACCCCATT-3’; 18S_R: 5’-CCATCCAATCGGTAGTAGCG-3’). Quant Studio 5 Real-Time PCR (Applied Biosystems, US) instrument was programmed for quantitation of C_t_ values for each gene.

### Methylation data processing

Whole-genome bisulfite sequencing (WGBS) data were generated from a total of six samples, comprising three post-stroke cognitive impairment (PSCI) and three post-stroke cognitively normal (PSCN) individuals. Genomic DNA from all samples was subjected to bisulfite treatment prior to sequencing. Raw paired-end sequencing reads were subjected to adapter trimming and quality control using fastp (v0.23.4). High-quality reads were aligned to the human reference genome of *Homo sapiens* (GRCh38, primary assembly) using Bismark (v0.22.3) with Bowtie2 (Li et al. 2009) (v2.5.4) as the underlying aligner, operating in paired-end mode with default parameters. PCR duplicates were removed using the Bismark deduplication module, and the resulting BAM files were sorted and indexed using Samtools (Li et al. 2009, Li et al. 2011) (v1.20). Cytosine methylation calling was performed using the Bismark methylation extractor, generating genome-wide cytosine methylation reports for downstream analyses.

### Differential Methylation Analysis

Differential DNA methylation analysis was performed between post-stroke cognitive impairment (PSCI) and post-stroke cognitively normal (PSCN) groups, with PSCN samples used as the reference group. CpG-level methylation data were derived from Bismark CpG cytosine reports generated for each sample and imported into the *methylKit* R package. CpG sites located at identical genomic coordinates on both DNA strands were merged to obtain strand-independent methylation estimates. Only CpG sites covered by a minimum of three sequencing reads were retained for downstream analysis. CpG sites were further filtered to retain those with non-zero methylation present in at least two samples per group, after which all qualifying CpG sites across samples were combined into a unified dataset using the *unite* function in *methylKit* (Akalin et al. 2012). Differential methylation at individual CpG sites was assessed using the chi-square test, as implemented by default in the *methylKit* package, which compares methylated and unmethylated read counts between PSCI and PSCN groups while accounting for sequencing coverage. CpG sites with a P value ≤ 0.05 were considered differentially methylated.

### Genomic regions of interest

Genomic regions encompassing *NTRK2* and selected regulatory elements were defined *a priori* to investigate CpG-level DNA methylation changes. Differentially methylated CpG sites were interrogated within these regions to assess potential regulatory and splicing-associated epigenetic mechanisms relevant to post-stroke cognitive impairment. Genomic coordinates were defined using the latest human genome assembly (GRCh38/hg38). The full-length *NTRK2* locus was defined as chr9:84,668,375–85,095,751. Genomic coordinates corresponding to the truncated form of NTRK2, as specified in this study, encompassed chr9:84,668,551–84,877,898. To evaluate regulatory influences, genomic regions corresponding to known transcriptional activators like *E2A*: chr19:1,609,291–1,652,615, *NeuroD1*: chr2:181,668,295–181,680,82 and *HIF1-alpha*: chr14:61,695,513–61,748,259; repressors like *HDAC2*: chr6:113,933,028–114,011,308, *MeCP*:chrX:154,021,573–154,137,103 and splicing-associated factors like *PRPF40B*: chr12:49,568,218–49,644,666 were also selected.

### Statistical Analysis

Hardy–Weinberg equilibrium (HWE) at the polymorphic sites was tested using a chi-square test with one degree of freedom. Genotype association was evaluated for p value, odds ratio and 95% confidence interval (CI) using Javastat (http://statpages.info/ctab2x2.html). Statistical analysis for gene expression data was per formed in Graph Pad Prism 5.0 using Mann–Whitney U-test to compare cases and controls. Two-tailed test was done with a significance level p < 0.05. Data presented as mean log_2_ transformed expression ± SEM. Correlation analyses between transcripts and raw score for cognitive parameters and GDS were performed by Spearman correlation analysis. Significantly differentially methylated CpG sites (P value ≤ 0.05) were extracted from these predefined genomic regions for region-specific epigenetic characterization, including analysis of CpG sites located within ±2 kb of transcription start sites (TSS). For each genomic region of interest, CpG sites were classified as hypermethylated or hypomethylated and summary statistics were computed, including the number of CpG sites, the sum of methylation differences, and the mean methylation difference.

## Results

### Demographic characteristics of study subjects

In the present study, demographic data and the principal variables for Ischaemic (IS) cases were represented for 152 cases with post stroke cognitive impairment (PSCI), 162 cases with good cognition (PSCN) and 11 age ethnicity-matched control individuals as tabulated in Table1. An older age of onset (P = 0.03797), female gender (P = 0.0474) and arterial involvement (PCA) showed (P = 0.0170) differential distribution between PSCI and PSCN cases. In corroboration with literature we also observed higher incidence of depression in PSCI group than PSCN in a statistically significant manner (P = 0.0047) (Table1).

### Gene expression of TRKβ isoforms and correlation with MEK2 mRNA expression and raw memory score

Gene expression of the full-length TRKβ^TK+^ was reduced in both memory preserved and memory impaired post stroke cases compared to controls by 22.54% and 23.15% [Mean log_2_ expression for TRKβ = −2.991±0.1046 (n=19); −3.005±0.1414 (n=21); −2.44±0.1388 (n=8), P=0.0273] respectively suggesting an overall lowering of full length receptor irrespective of cognitive status [Figure 1A]. In contrast, mRNA level of truncated TRKβ-SHC was similar between controls and memory preserved cases but lowered in memory impaired groups from former ones by 13.5% and 17.985% respectively [Mean log_2_ expression for TRKβ-SHC= - 2.378±0.1564 (n=8); −2.268±0.1879 (n=19); −2.75±0.09982 (n=21), P_1ANOVA_=0.0355, P_2_ = 0.0168] indicating neuroprotective role of this isoform in memory preserving among post stroke survivors [Figure 1B]. However, truncated TRKβ–T1 expression did not show any differential expression [Figure 1C] among the groups [Mean log_2_ expression for TRKβ-T1= − 2.397±0.1250 (n=8); −2.968±0.1415 (n=19); −2.776±0.0.1157 (n=21), P_1ANOVA_=0.0540]. Our further intra group comparative analysis of TRKβ isoforms identified alteration in stoichiometry between the transcripts along with significant overexpression of TRKβ-SHC among MP survivors [P_ANOVA_ = 0.0085] [ Figure 1E].

**Figure 1:**
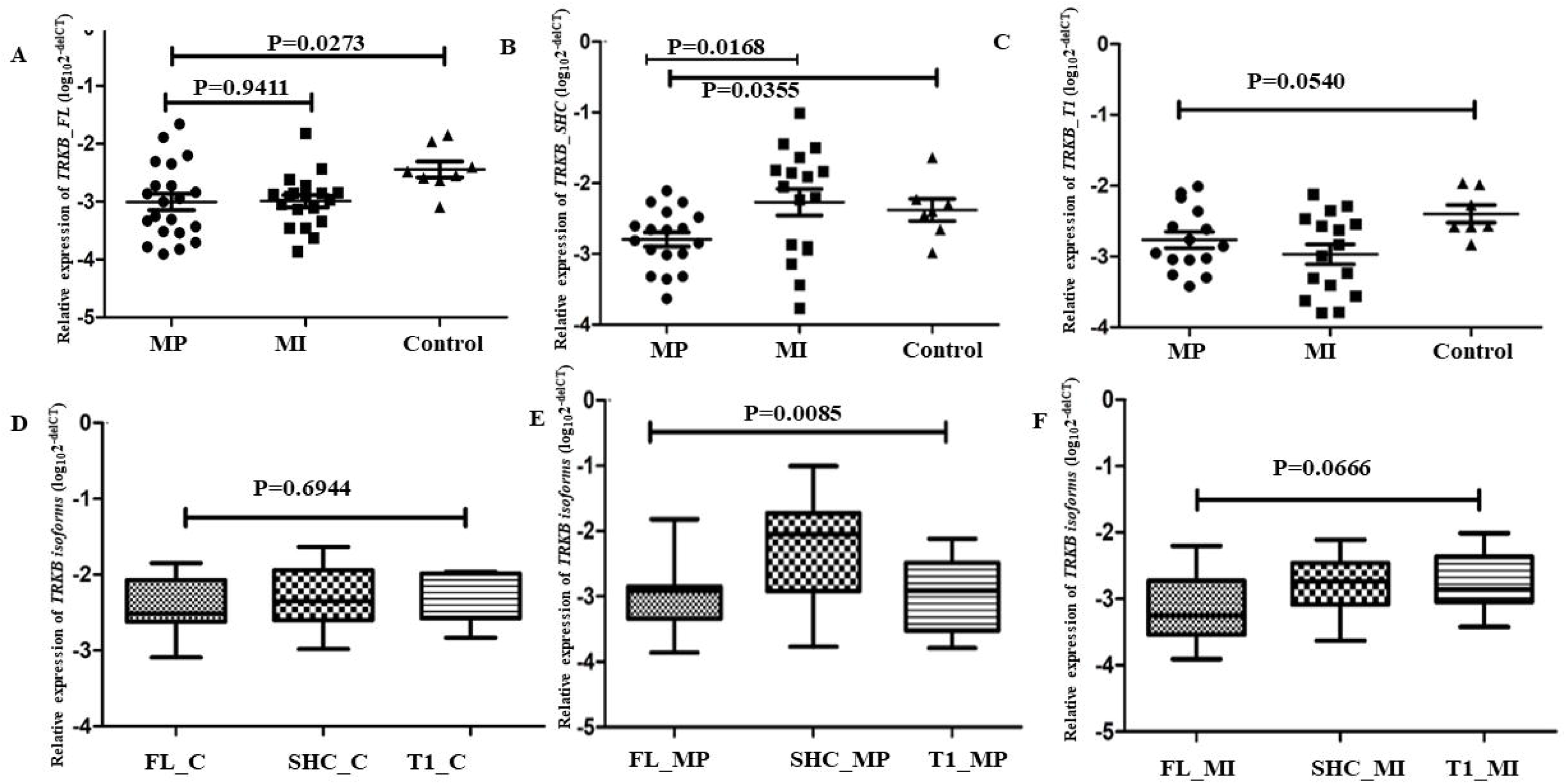
Comparative analysis of three isoforms of TRKβ among post stroke survivors with different memory status. **(A).** Quantification of TRKβ_FL mRNA levels in human PBMC from controls (n = 8) and Post-stroke memory impaired (MI) patients (n = 21), Post-stroke memory preserved (MP) patients (n = 19) normalized to the house keeping gene 18S using ANNOVA and t-test (P_A_ = 0.0273; P = 0.9411). **(B).** Determination of TRKβ_SHC levels among the said groups (P_A_ = 0.01683; P = 0.0355) **(C).** Quantification of TRKβ_T1 mRNA levels in same individuals (P_A_ = 0.0540). **(D. E. and F).** Intragroup Relative expression of TRKβ isoforms levels among controls (C), memory preserved (MP) and memory impaired (MI) subjects respectively (P_A_ = 0.6944, 0.0085 and 0.0666). Data presented as mean ± standard deviation in all graphs. The significance level was set at P ≤ 0.05

Owing to the complex yet important role of MEK signalling in post stroke recovery and cognition, as well post translation regulation of SHC on MEK activation, next, we performed a correlation study between TRKβ-SHC, MEK gene expression and raw memory score in a study cohort of n=35 irrespective of disease phenotype. Our finding showing positive a statistical significant correlation of TRKβ-SHC with MEK expression level and memory score [r = 0.5814, P = 0.0005 (95% CI = 0.2819-0.7776); r = 0.5441, P= 0.0005 (95% CI= 0.2578-0.7425)] respectively further validated neuroprotective effect of TRβ –SHC for post stroke cognitive function [Fig 2A-2B].

**Figure 2:**
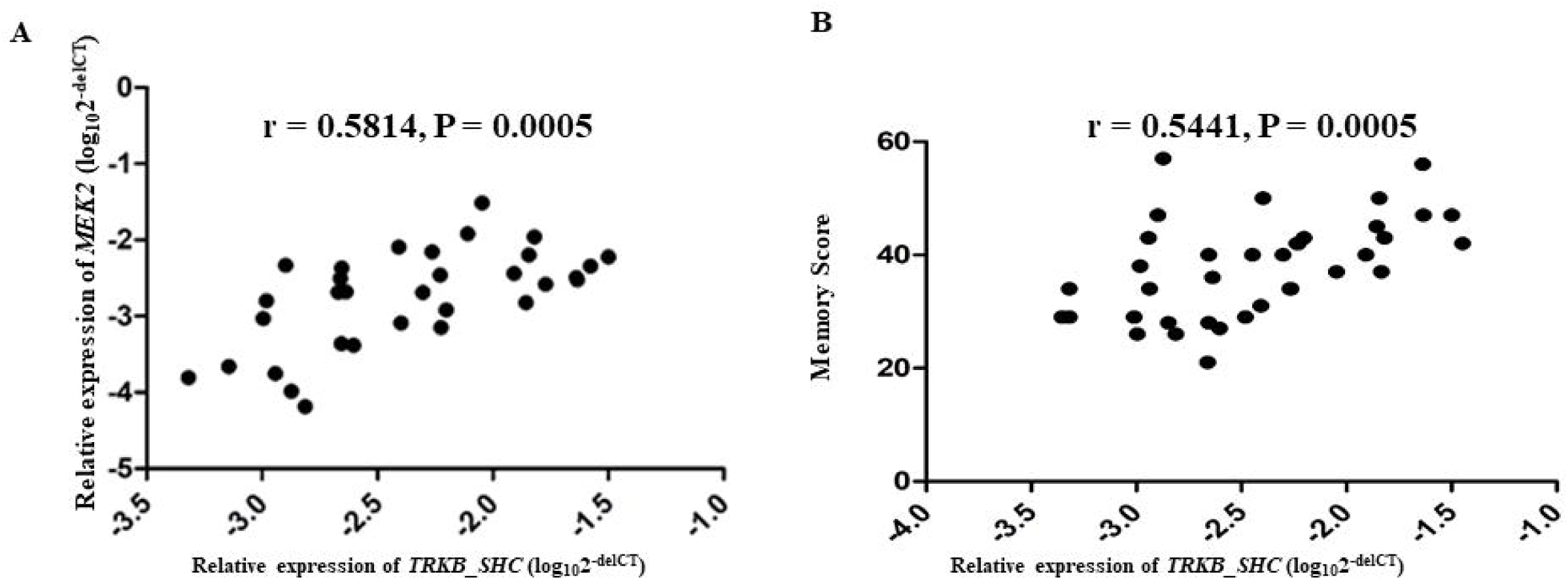
**Significant positive correlation of TRK**β**-SHC expression with** (**A)** MEK2 expression and (**B)** Memory scores. Diagrammatic representation showing putative differential role of TRKβ-FL and TRKβ-SHC in post stroke memory outcome.

### Selection of SNV in TRKβ-SHC specific rs6559833T/C for association study with Post-stroke cognitive impairment

Since miRNAs and miRNA binding sites are potent regulator of gene expression, here we aimed to identify putative functional SNV specific to TRKβ-SHC transcript conferring PSCI susceptibility among Eastern Indian stroke patients. For such *in-silico* prediction TRKβ-SHC was scanned through Target Scan_7.1 (http://www.targetscan.org/vert_71/) and miR Search v3.0 (https://www.exiqon.com/miRSearch) were used. Both prediction algorithms predicted hsa-miR-6770-3p to be a putative target for rs6559833, a SNV at 3’UTR+5295 which was not investigated before. The 8-mer seed match between hsa-miR-6559833-5p and the SNV had a context score of binding of 0.71 when the C-but no binding predicted for the T-allele of the same.

### Distribution of TRKβ genotypes

In the present study, 152 PSCI and 162 PSCN patients were initially recruited for genotype analysis. The genotype and allelic frequencies of this variant among both the group were in Hardy–Weinberg equilibrium). However, both genotype and allele frequencies of rs6559833 showed statistically significant differences between PSCI and PSCN group (P=0.0442 & 0.0027) [Table 2]. For the rs6559833 ‘T-allele’ [OR=1.437; 95% CI=1.009-2.045; P=0.0442] and ‘TT genotype’ [OR=1.755; 95%CI=1.066-2.891; P=0.0027] were observed to be associated with risk for PSCI.

**Table 2:** Distribution of genotype and allele frequencies of *TRK*β *(rs6559833)* variant among Stroke patients and Controls.

| Group | Genotype frequency (%) |  |  | Allele frequency (%) |  | P-value | Odds ratio (95% CI) |
| --- | --- | --- | --- | --- | --- | --- | --- |
|  | TT | TC | CC | T | C |  |  |
| rs6559833 |  |  |  |  |  |  |  |
| Memory Impaired (n = 136) | 62 (45%) | 58 (43%) | 16 (12%) | 182 (67%) | 90 (33%) | T vs C: **0.0442** | 1.437 (1.009 – 2.045) |
| Memory Preserved (n = 130) | 42 (32%) | 68 (52%) | 20 (16%) | 152 (58.5%) | 108 (41.5%) | TT vs CC+CT: **0.0027** | 1.755 (1.066 – 2.891) |
| Depressed (n = 152) | 64 (42%) | 64 (42%) | 24 (16%) | 192 (63.2%) | 112 (36.8%) | T vs C: 0.5682 | 0.9053 (0.643 – 1.274) |
| Non-depressed (n = 136) | 54 (40%) | 70 (51%) | 12 (9%) | 178 (65.5%) | 94 (34.5%) | TT vs CC+CT: 0.6794 | 1.104 (0.689 – 1.768) |

GDS ≥ 9: Depressed; GDS <9: Non-depressed

### Association of rs6559833 with cognitive, depression raw scores and TRKβ expression

An overall positive association among ‘TT’ carriers of the variant only for BMSE score represented mild genotypic association with overall cognition deterioration [Mean score 23.33±5.52 vs 25.18±4.93, P = 0.04881] [Table 3]. However, it failed to show further association with raw scores of different subdomains of cognition like memory, language and GDS. Next, to investigate whether TRKβ gene expression can be linked to rs6559833T/C, irrespective of disease phenotype, the minor allele carrying genotypes were grouped together. By doing this, we found that no difference in mean gene expression between the ‘C’ allele carrier and ‘T’ allele of the same (data not shown).

**Table 3:**
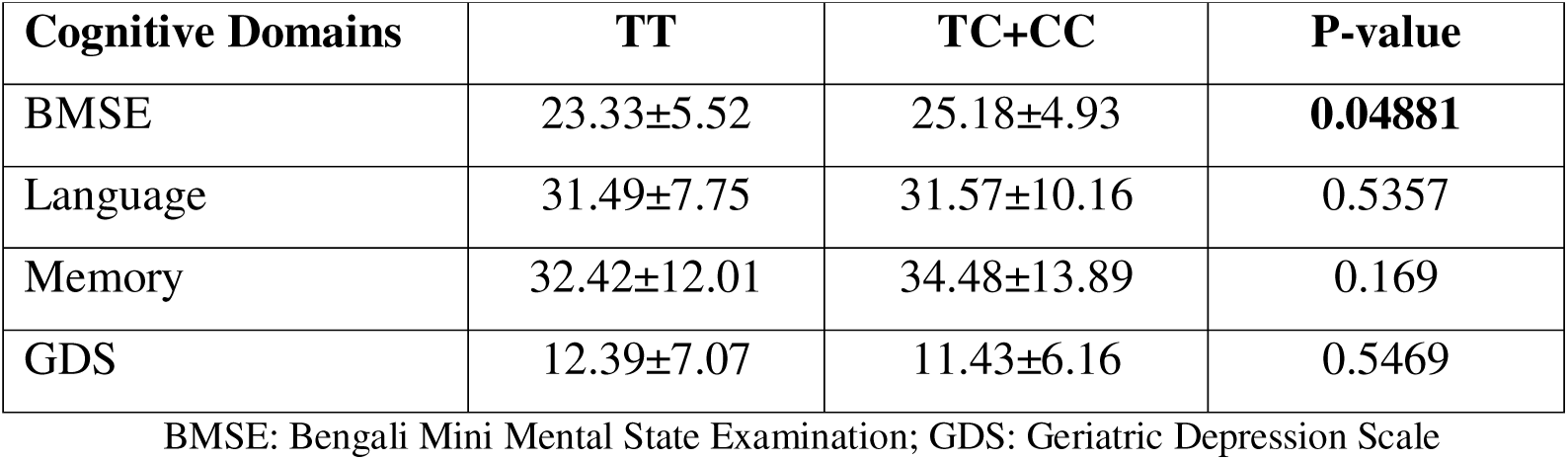
Comparison of cognitive parameters among the Post-Stroke survivors according to their rs6559833 genotypes.

| **Cognitive Domains** | **TT** | **TC+CC** | **P-value** |
| --- | --- | --- | --- |
| BMSE | 23.33±5.52 | 25.18±4.93 | **0.04881** |
| Language | 31.49±7.75 | 31.57±10.16 | 0.5357 |
| Memory | 32.42±12.01 | 34.48±13.89 | 0.169 |
| GDS | 12.39±7.07 | 11.43±6.16 | 0.5469 |

BMSE: Bengali Mini Mental State Examination; GDS: Geriatric Depression Scale

### Region-Specific DNA Methylation Patterns across the *NTRK2* Locus and its transcriptional regulators

With the knowledge of regulatory role of DNA methylation on gene expression we quantified differentially methylated CpG sites (DMCs) across the *NTRK2* locus, including the full-length gene body, a truncated genomic region, and the promoter-proximal transcription start site (TSS ±2 kb) (Figure 3A–F). Distinct region-specific differences were observed in both the distribution and magnitude of methylation changes. Across the full-length NTRK2 locus, hypermethylation predominated (Figure 3A). A total of 71 CpG sites were hypermethylated, compared to 9 hypomethylated sites. Hypermethylated CpGs showed a large cumulative methylation increase (sum meth.diff = 1591.85) and a high mean methylation difference (mean meth.diff = 22.42), whereas hypomethylated CpGs exhibited smaller cumulative differences in methylation (sum meth.diff = −72.22; mean meth.diff = −8.02). Analysis of the truncated *NTRK2* region showed a reduced number of DMCs while maintaining the same directional bias (Figure 3C-D). Nineteen CpGs were hypermethylated (sum meth.diff = 433.47; mean meth.diff = 22.81), and nine CpGs were hypomethylated (sum meth.diff = −72.22; mean meth.diff = −8.02).

**Figure 3:**
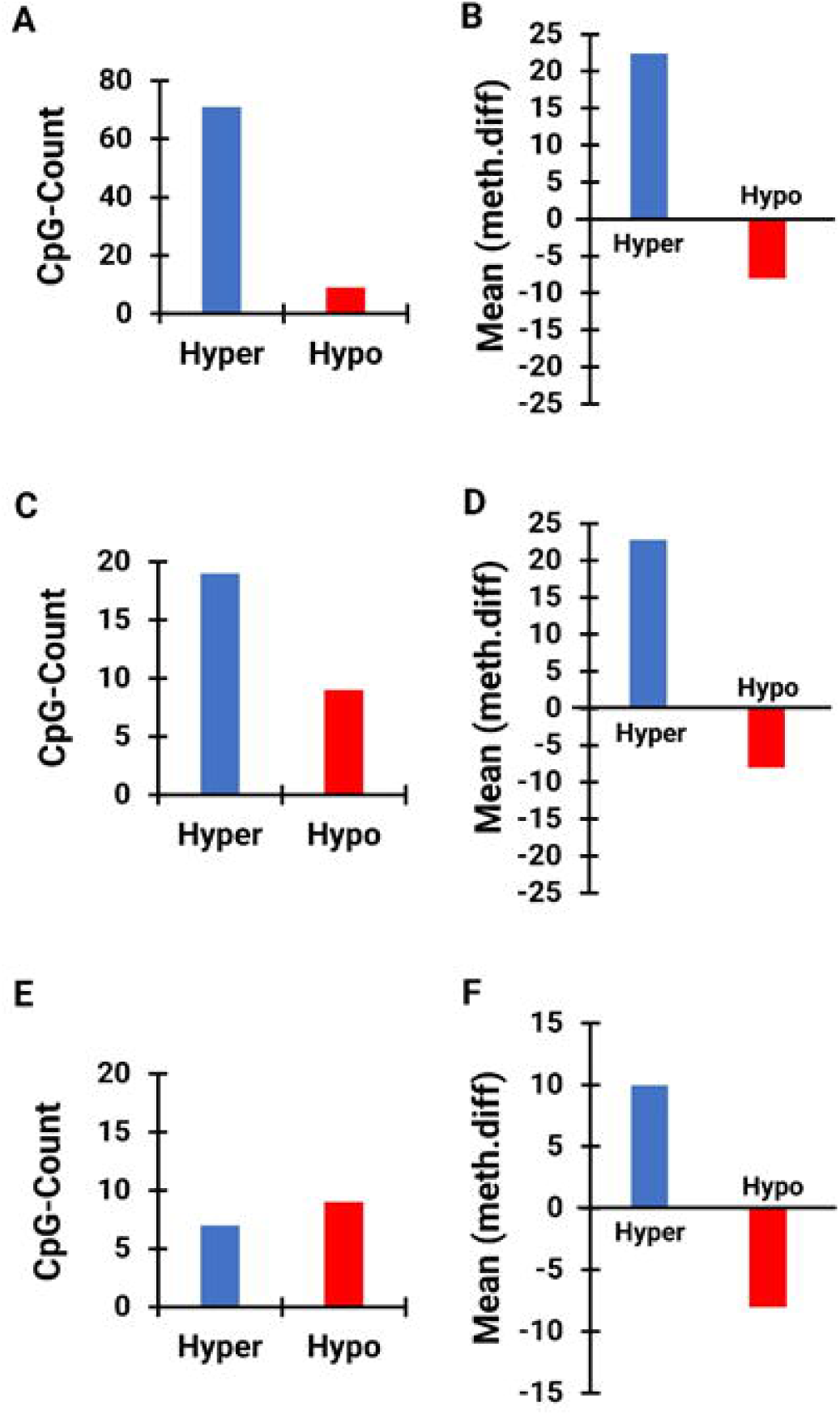
Differential DNA methylation patterns across the *NTRK2* Gene. (A–B) Full-length *NTRK2* Gene. **(A)** Bar plot showing the number of differentially methylated CpG sites categorized as hypermethylated (blue) and hypomethylated (red). **(B)** Mean methylation difference (meth.diff) for hyper and hypomethylated CpGs within the full-length *NTRK2* region. **(C–D) Truncated *NTRK2* Gene. (C)** Distribution of hyper and hypomethylated CpG sites within the truncated *NTRK2* Gene. **(D)** Corresponding mean methylation difference for hyper and hypomethylated CpGs in the truncated Gene. **(E–F) Promoter-proximal region (TSS ±2000 bp). (E)** Number of differentially methylated CpG sites at the transcription start site (TSS ± 2 Kb) region of *NTRK2*. **(F)** Mean methylation difference for hyper and hypomethylated CpGs within the TSS-proximal region.

In contrast, the TSS-proximal region (±2 kb) displayed a more balanced methylation profile (Figure 3E, F). Seven CpGs were hypermethylated (sum meth.diff = 69.72; mean meth.diff = 9.96), while nine CpGs were hypomethylated (sum meth.diff = −72.22; mean meth.diff = −8.02). Compared with the gene body, methylation differences at the TSS were of lower magnitude.

To further characterize regulatory context, DMCs overlapping known transcriptional activators (E2A, HIF1-α, NeuroD1), repressors (HDAC2, MeCP), and the splicing factor PRPF40B were analyzed (Figure S1–S2, Figure 4i-4iv). Activator-associated regions showed a predominance of hypermethylated CpGs within gene-associated intervals *(e.g.,* E2A: 181 hyper vs 5 hypo CpGs; mean meth.diff = 16.91), while promoter-associated CpGs were limited or absent for several factors. Repressor-associated regions displayed strong gene-associated hypermethylation (e.g., MeCP: mean meth.diff = 36.19). PRPF40B-associated regions showed substantial gene-associated hypermethylation, with 50 hypermethylated CpGs within the gene body (sum meth.diff = 1094.33; mean meth.diff = 21.89) compared to only 4 hypomethylated CpGs (sum meth.diff = −57.60; mean meth.diff = −14.40), whereas the TSS-proximal region contained markedly fewer CpGs (2 hypermethylated and 1 hypomethylated site).

**Figure 4:**
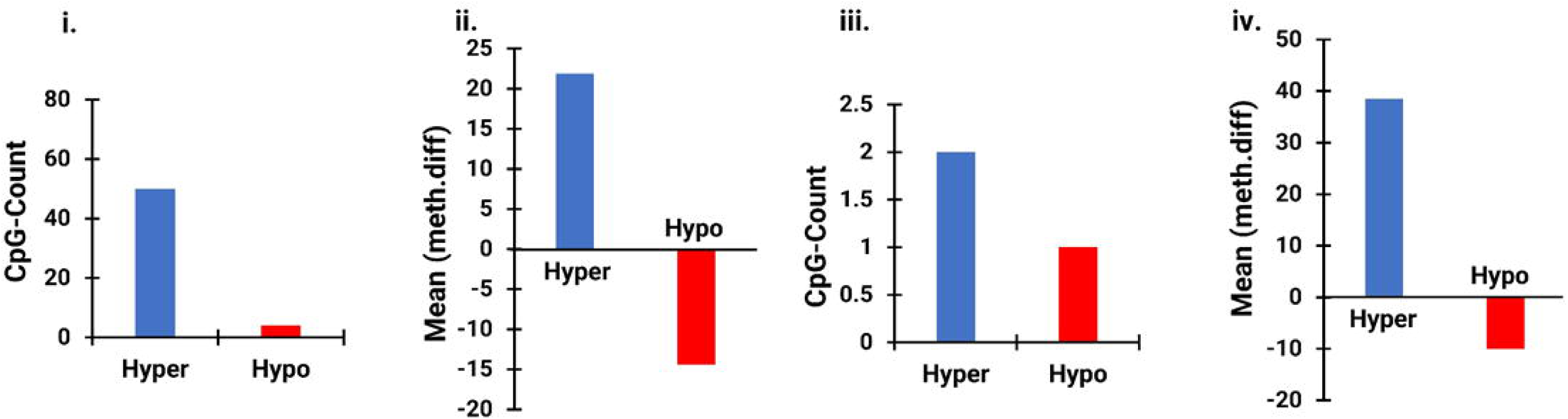
PRPF40B Splicing factor. (i–ii) CpG count (i) and mean methylation difference (ii) for differentially methylated CpGs across the **full-length NTRK2 locus** overlapping PRPF40B-associated regions. (iii–iv) Corresponding CpG counts (iii) and mean methylation differences (iv) for **TSS-proximal regions (±2 kb)** associated with PRPF40B.

## Discussion

Given that the occurrence of PSCI is a complex process, it is affected by many factors such as genetic susceptibility, the patient’s age, stroke subtype, specific location of the lesion, comorbidities, emotional state, sociodemographic factors, etc. However, many allied studies have emphasized on role of BDNF in patient derived studies than TRKβ. Although, an imbalance of Trkβ-FL/Trkβ-T1 was reported in neurons of infarcted human brain and rat neurons that were subjected to in vivo or in vitro excitotoxicity (Vidaurre et al. 2012), unlike Schizophrenia and Alzheimer’s disease, the role of TRKβ-SHC in post stroke cognition was less characterised (Wong et al. 2013, Ansaloni et al. 2011). Therefore, the purpose and significance of this study is a) to explore the role of TRKβ-SHC in post stroke cognition through transcript analysis in human PBMC mimicking lesion site, and b) look for potential contribution of DNA methylation in TRKβ gene and isoform specific 3’UTR SNV towards alteration in isoform expression pattern.

Towards achieving our objectives, our major findings include, an over expression of TRKβ-SHC isoform but not for TRKβ-FL and TRKβ-T1 in PBMC of PSCN patients than PSCI patients, followed by its positive correlation with MEK mRNA level, suggesting a neuroprotective role TRKβ-SHC towards better learning and memory pathway among post stroke survivors. An altered ratio of TRKβ-SHC and TRKβ-FL between MP (1:0.758) and MI (1:915) also directed alternative splicing of TRKβ as a putative player for differential outcome in post stroke recovery phase. Owing to the fact stating dual role of MEK2 signaling pathway in post stroke phases, our positive correlation between TRKβ-SHC and MEK2 expression level further reconfirms neuroprotective nature of truncated version of TRKβ as well propose as putative therapeutic target for cognitive improvements among post stroke survivors.

The similar observation like increased mRNA expression for truncated TRKβ receptor isoform was observed in prefrontal cortex of people with schizophrenia (Wong et al. 2013. However, this overexpression was linked to disease pathogenesis through impairments in inhibitory transmission and cortical plasticity in them. On the other hand, in case of AD TRKβ-SHC overexpression was found to play neuroprotective role by dampening BDNF-stimulated TRKβ -TK second messenger signalling via the MEK pathway but not the AKT pathway to promote cell survival without activating an already overactive MEK signalling pathway in AD (Ansaloni et al. 2011).

A previous study describing the miRNA mediated regulation of TRKβ-SHC expression (Wong et al. 2014), made us interested to determine the role of a 3’UTR variant (rs6599833C/T) of it showing putative miRNA binding site. Here, we have identified a significant association between rs659983 and BMSE score representing cognitive status of post stroke survivors from Eastern India. In contrast, absence of such association with cognitive subdomains was a major reason for not performing additional *in-vitro* study. The inconsistency between prediction and observed association can be explained by population size and or selection of ethnic background.

Like genetic variants, DNA methylation can significantly affect the isoform formation by regulating alternative splicing and regulating transcription from alternative transcription start site, here we have compared the methylation profile of NTRK2 along with its activators repressors and splicing factor across their promoter and genic region between PSCI and PSCN cases. Concordant to the literature, for each of gene we studied here using PBMC showed less methylation mark than gene-body methylation. Although promoter region of NTRK2 gene did not show differences between the groups, an overall hypermethylation was observed in gene body of NTRK2 encoding longest transcript as well encoding TRKβ-SHC isoform. Thus, gene body methylation in NTRK2 gene may be a reason for less expression of neuroprotective truncated one as genic methylation is often correlated with little or no transcription from alternative internal promoters bringing variation in isoform’s quantity. Likewise, it may also be also influenced by hypermethylation status of PRPF40B, a NTRK2 specific splicing factor (Duarte-Ruiz et al. 2025) [Fig 5A-B]. Unlike promoter methylation gene body methylation often results into higher gene expression. However, although both the activators and repressors of NTRK2 also showed similar hypermethylation in PSCI cases, but it is hard to correlate as these can regulate expression of multiple genes at a same time. Similar observation was also reported by a recent study, describing involvement of epigenetic modifications of *NTRK2* in memory modulation and conferring risk and symptoms of PTSD in case of traumatic experiences (Vukojevic et al. 2020).

**Figure 5:**
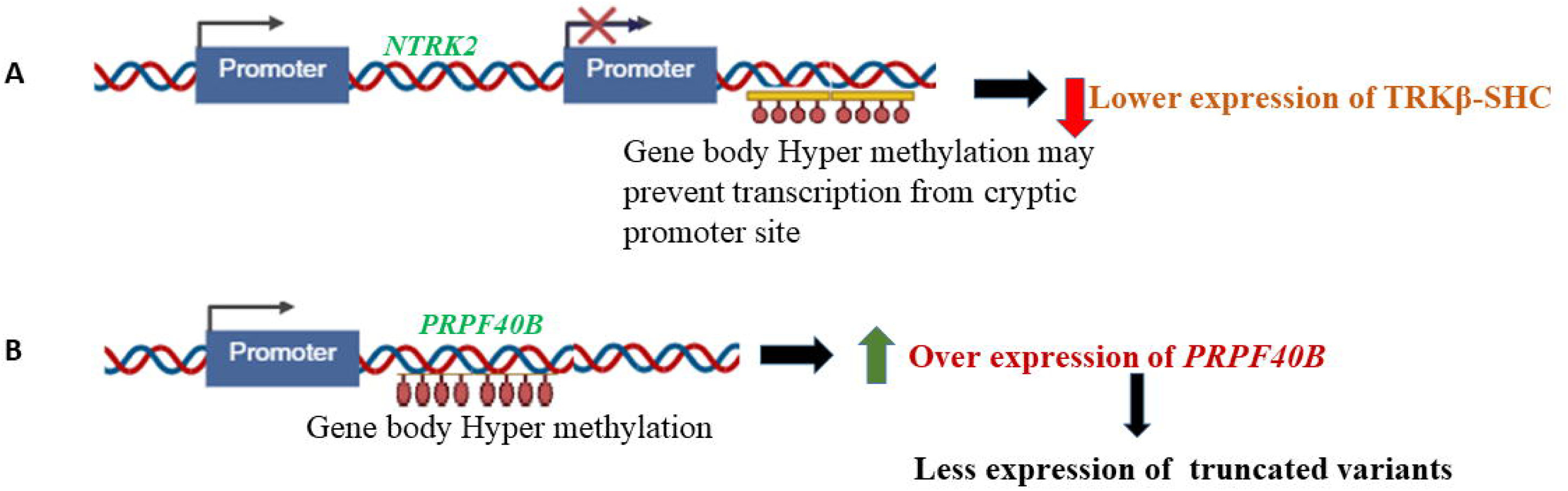
Schematic representation of the proposed regulatory effects of gene body methylation in (A) **NTRK2** and (B) **PRPF40B** on TRKβ–SHC mRNA expression.

Despite our findings, the limitation of our study includes a) use of PBMC as biosource while PBMC infiltration partially mimics the CNS environment, it does not accurately reflect CNS-specific regulatory mechanisms; b) no supporting evidence from protein expression data; c) the small sample size for methylation assay. Therefore, future studies using human post mortem brain as biosource for correlation analysis between RNA and protein level, identification of TRKβ-SHC isoform-specific interactome and deciphering molecular factors influencing alternative splicing may constitute therapeutic targets for post stroke cognition.

Therefore, in summary, considering our present study on TRKβ and previous study describing lower plasma BDBF level and lower BDNF gene expression in PSCI than PSCN, we may suggests two different scenario for these differential outcomes. The reduced BDNF and TRKβ -FL along with TRKβ-SHC transcript converge to memory impairment by restricting MEK signalling required for synaptic plasticity in post stroke recovery phase. In contrast, in case of PSCN, higher level of BDNF but lesser TRKβ-FL may be compensated by upregulation of TRKβ-SHC and formation of more -SHC homodimers and FL/SHC heterodimers. These inactive receptor complex may sequester excess BDNF and thus restore ligand receptor ratio to prevent excitotoxicity as well maintaining the optimum BDNF-TRKβ ratio required for better cognitive performance through MEK signalling [Fig 6]. On the other hand, from the findings describing similar ratio between TRKβ-SHC: TRKβ-FL in case of healthy controls and memory impaired ones (1:0.975, 1:0.915) but different from memory preserved ones (1:0.758), we can also propose that rather than ratio but the cumulative effect of relative expression of NTRK2 isoform expression and BDNF level plays an important role post stroke memory status

**Figure 6:**
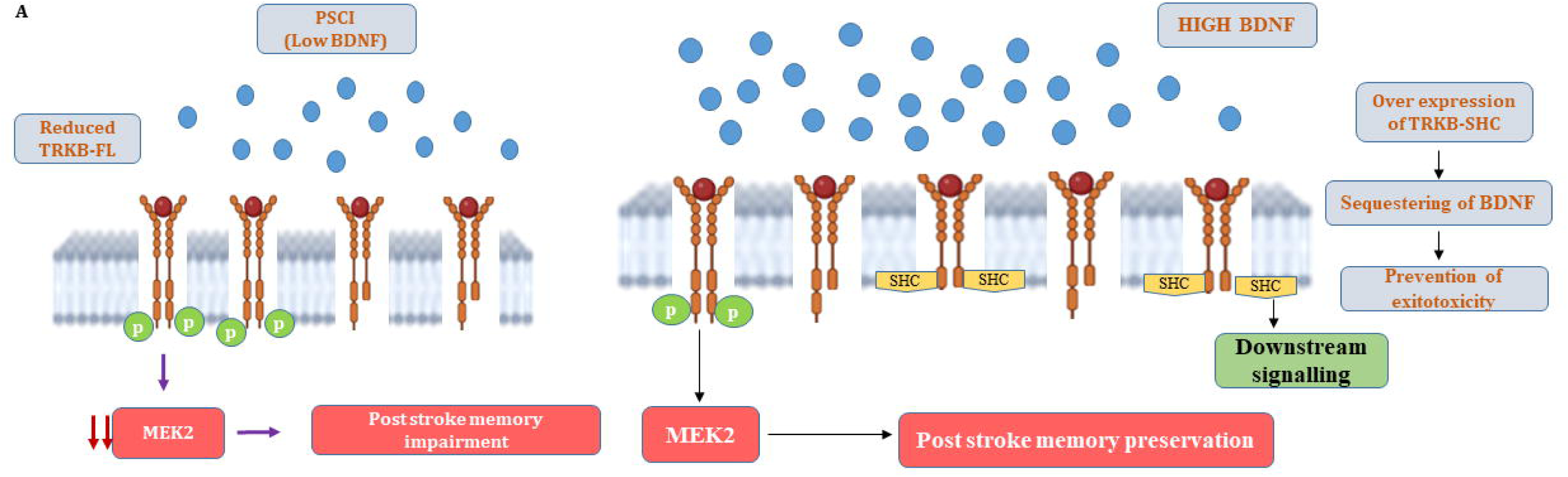
Schematic illustration of TRKβ–SHC–mediated neuroprotection contributing to post-stroke memory preservation.

**Figure S1. Differential DNA methylation patterns at transcriptional activators of the *NTRK2* Gene.**

**(A) E2A Transcriptional Factor**. (i) Bar plots representing the number of differentially methylated CpG sites and the mean methylation difference (meth.diff) (ii) within the **full-length *NTRK2* gene** associated with E2A binding regions, categorized as hypermethylated (blue) and hypomethylated (red). (iii–iv) Corresponding CpG counts (iii) and mean methylation differences (iv) for **TSS-proximal regions (±2 kb)** associated with E2A.

**(B) HIF-1**α **Transcriptional Factor.** (i–ii) Distribution of hyper and hypomethylated CpG sites (i) and their mean methylation differences (ii) across the **full-length *NTRK2* locus** overlapping HIF-1α–associated regions. (iii–iv) CpG counts (iii) and mean methylation differences (iv) for **TSS-proximal regions (±2 kb)** associated with HIF-1α.

**(C) NeuroD1-associated regions.** (i–ii) CpG count (i) and mean methylation difference (ii) for differentially methylated CpGs within the **full-length *NTRK2* locus** overlapping NeuroD1-associated regions. No NeuroD1-associated CpGs were detected within the TSS-proximal region under the applied criteria.

**Figure S2. Differential DNA methylation patterns at repressor of the *NTRK2* Gene.**

**(A) HDAC2 Transcriptional Factor (repressor).** (i–ii) Number of hyper- and hypomethylated CpG sites (i) and mean methylation difference (meth.diff) (ii) within the **full-length NTRK2 locus** overlapping HDAC2-associated regions. (iii–iv) Corresponding CpG counts (iii) and mean methylation differences (iv) for **TSS-proximal regions (±2 kb)** associated with HDAC2.

**(B) MeCP Transcriptional Factor (repressor).** (i–ii) Distribution of differentially methylated CpG sites (i) and their mean methylation differences (ii) across the **full-length NTRK2 gene** overlapping MeCP-associated regions. (iii–iv) CpG counts (iii) and mean methylation differences (iv) for **TSS-proximal regions (±2 kb)** associated with MeCP.

## Data Availability

The datasets generated and/or analyzed during the current study are not publicly available due to ongoing parallel studies involving novel observations, as well as planned future publications arising from downstream analyses of these datasets. However, the data are available from the corresponding author upon reasonable request.

## Acknowledgements

The authors are thankful to the patients, family members and healthy individuals who participated in the study.

## Author Contributions

DS contributed towards project administration, methodology, funding acquisition, writing – original draft, and experimental work and data analysis; AB contributed towards project administration, methodology, funding acquisition, writing–original draft, and experimental work and data analysis; RC contributed methylation data analysis, manuscript writing; SC contributed towards methylation data analysis, manuscript writing, providing infrastructural facilities for data analysis; AR, SR, SM contributed towards experimental work and data analysis; TKB, KCG & JM contributed towards patient recruitment, clinical evaluation and manuscript review & editing; SPH contributed towards manuscript review & editing and providing infrastructural facilities for experimental work. All authors reviewed the manuscript.

## Declarations

### Ethical Approval

All procedures performed in studies involving human participants were in accordance with the ethical standards of the “Institutional Ethics Committee, Nil Ratan Sircar Medical College, Kolkata”. The Institutional Ethics Committee, Nil Ratan Sircar Medical College, Kolkata, approved the study protocol (Memo No: NRSMC/IEC/89/2021, Date: 23.12.2021 and NRSMC/IEC/038/2025, Date: 07.03.2025). Informed consent was taken as per the guidelines of the Indian Council of Medical Research, National Ethical Guidelines for Biomedical and Health Research involving human participants, India.

### Consent to Participate

All procedures performed in studies involving human participants were in accordance with the ethical standards of the participating institutions. The informed consent form was taken either from patients or caregivers.

