## Supplementary figures and images for "Transcriptional alteration in TRKβ-SHC isoform as a neuroprotective factor for post stroke memory outcome"

### Supplement Fig 1

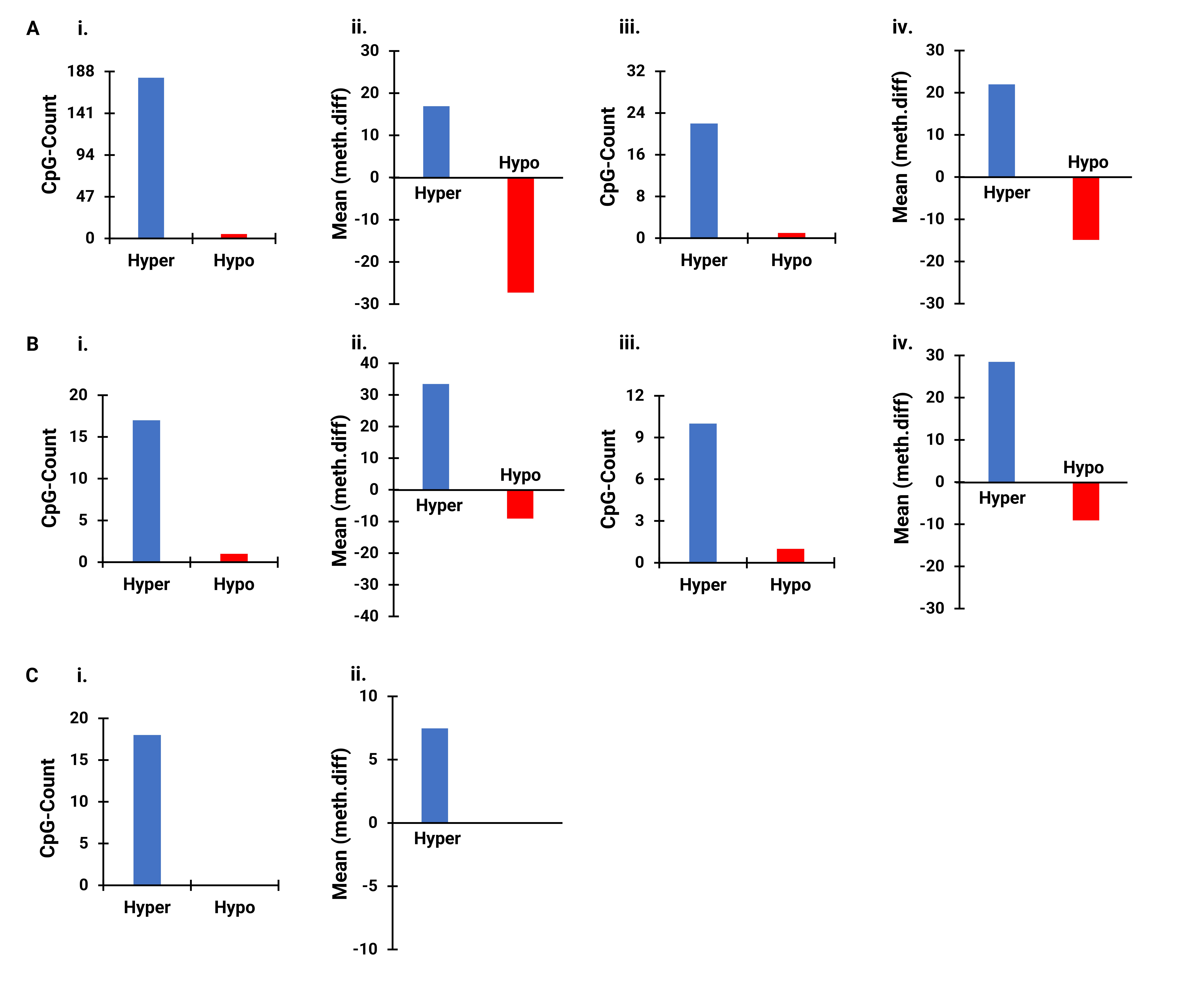

### Supplement Fig 2

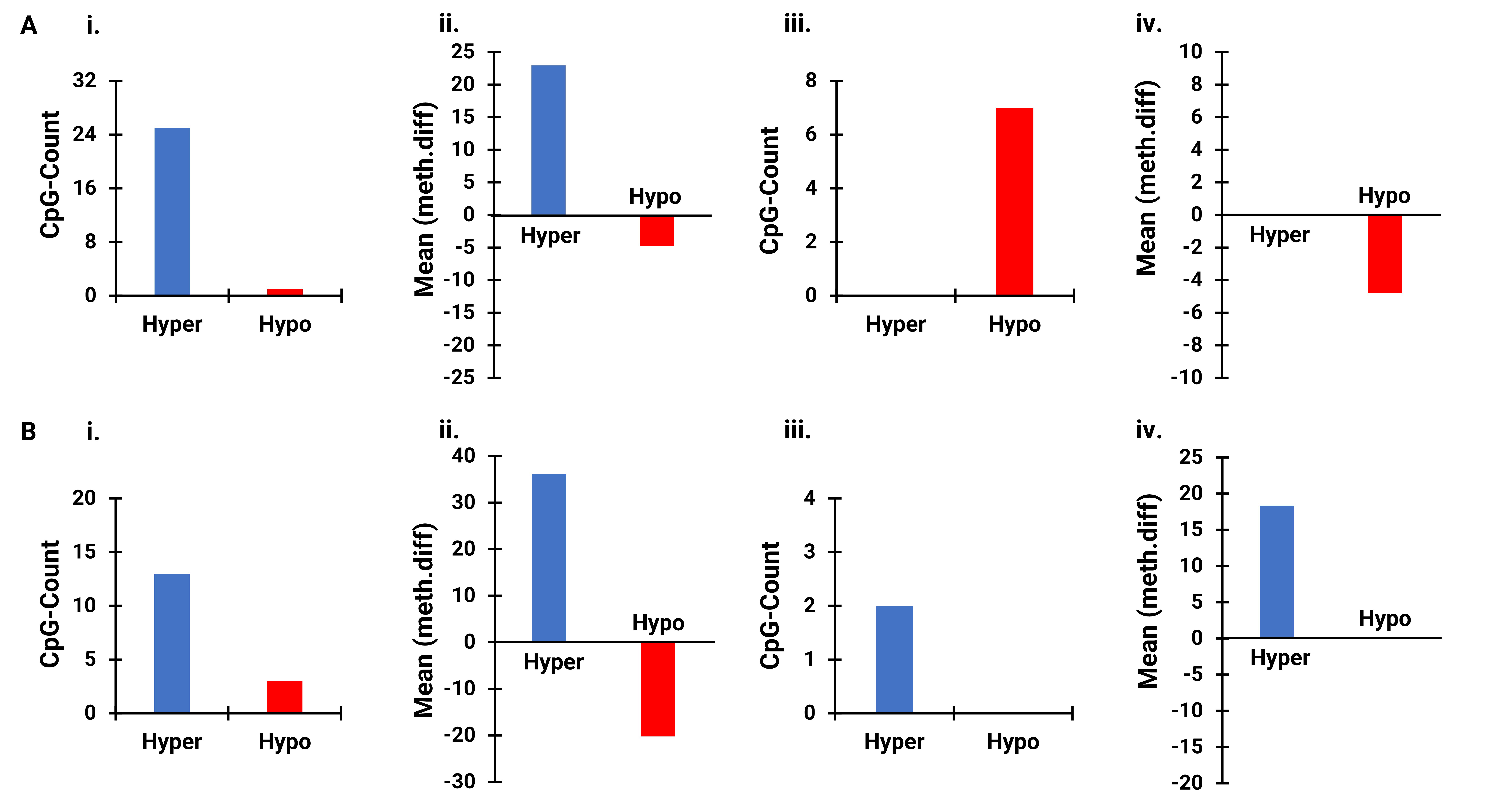
